# Field Evaluation of a Low-cost, Simplified Method to Detect and Characterize Toxigenic *V. cholerae* O1 from Environmental Water in Lagos, Nigeria

**DOI:** 10.64898/2026.10.23.26366021

**Authors:** Akinsinde Kehinde Adewale, Diana Dinowitz, David A. Sack, Wensheng Luo, Nathaniel Matteson, Kirabo Tess Ayazika, Mena Kazerounian, Bamidele Tajudeen Akanji, Amanda K Debes, Shirlee Wohl, Iwalokun Bamidele Abiodun

## Abstract

Cholera is caused by toxigenic *Vibrio cholerae* serogroups O1 or O139 and is transmitted through contaminated water or food. Conventional detection methods for *V. cholerae* in environmental water samples, including membrane filtration and culture, typically require about three days, specialized equipment, and technical expertise. A simple and rapid method for detecting the *V. cholerae* O1 in water could support outbreak prevention, environmental surveillance, and studies of persistence and transmission. Here, we describe a low-cost, streamlined method for detecting toxigenic *V. cholerae* O1 in environmental water which can detect *V. cholerae* O1 within 24 hours. We filter one liter of the water through gauze, incubate the gauze in alkaline peptone water (APW) overnight at 37 C, and test the APW sample with a serogroup O1-specific rapid diagnostic test (RDT). To preserve nucleic acid material from APW broth, we place two drops on each circle of a filter paper (Whatman 903), dry it, and later analyze the DNA using PCR and whole genome sequencing. We evaluated this method using 95 environmental samples collected from 17 sites in Lagos, Nigeria. 54 samples were RDT positive and each of these samples were confirmed by culture and by PCR targeting *ompW* and *ctxA* as toxigenic *V. cholerae* O1. Whole-genome sequencing identified these isolates as belonging to the AFR12 lineage.

Among the RDT-negative samples, PCR detected 24 samples positive for *ompW* but negative for *ctxA*, consistent with non-toxigenic non-O1 *V. cholerae*, and 11 of these were further identified as non- toxigenic serogroup O139 by qPCR using a commercial assay (Bioperfectus). The cost for the RDT test was <$2.50 per water sample. This low-cost APW enrichment and RDT-based approach enables identification of environmental water contaminated with toxigenic *V. cholerae* O1 within 24 hours and enables the collection of samples useful for further bacterial molecular characterization.

**Author Summary:** Standard methods to identify water samples contaminated with *V. cholerae* serotype O1 are commonly used, but their use for public health practice has been limited due cost, time, and resource requirements. This study describes a convenient, efficient and inexpensive method to detect *V. cholerae* O1 from environmental water samples and evaluates this method using water samples collected from 17 sites in Lagos, Nigeria during 2025. The method involves filtering one liter of water through a gauze pad, followed by overnight incubation of the gauze in alkaline peptone water (APW) enrichment broth. The APW is then tested for cholera using a rapid diagnostic test (RDT) specific for *V. cholerae* O1. Among 95 environmental water samples, 54 were positive for *V. cholerae* O1 using the RDTs. This method provides results within one day at a cost of <$2.50 USD per sample. If there is need for additional studies, RDT positive samples can be sent for culture, and nucleic acid from the APW can be saved by spotting the APW on filter paper. The filter paper can then be stored at room temperature indefinitely. All of the RDT positive samples were confirmed by culture and PCR Among the PCR negative samples, 24 samples were PCR positive for non-O1 *V. cholerae,* including 11 that were positive for non-toxigenic *V. cholerae* O139 by qPCR. Whole genome sequencing of the samples found the RDT positive samples to be of the AFR12 lineage, and the DNA contained many genetic markers of antibiotic resistance. Identifying cholera contaminated water quickly provides information to health authorities which can facilitate interventions to reduce transmission and can also provide samples for further epidemiologic and molecular studies.

## Introduction

Cholera is an acute waterborne diarrheal disease caused by toxigenic *Vibrio cholerae* serogroups O1 or O139 and can become fatal within hours if left untreated.^1^ Death are due to profuse watery diarrhea which results in rapid fluid and electrolyte loss leading to severe dehydration, metabolic acidosis, and shock. Early recognition of cases is therefore essential to enable timely clinical management including replacement of fluids to correct the dehydration, and antibiotics to kill the bacterial pathogen. When outbreaks occur, prompt public health interventions can limit its spread. Rapid diagnostic tests (RDTs) are increasingly available to detect *V. cholerae* O1 in stool samples to support early case detection.^2,3^ Stools with a positive RDT can then be confirmed by culture or PCR,^4^ helping to trigger an appropriate outbreak response.

Cholera occurs when a susceptible person ingests water or food contaminated with toxigenic *V. cholerae* O1. In turn, the feces of cholera patients can contaminate environmental water in areas with poor water and sanitation. Through this contamination, the disease can spread from patients to other people who use the water for drinking or food preparation. Thus, during outbreaks, wastewater or other water bodies can become contaminated with *V. cholerae* O1 and be the source for onward transmission. Although *V. cholerae* can often be detected in environmental water, among more than two hundred serotypes, only serotypes O1 and O139 are able to cause epidemic cholera; in fact, in recent years, only serotype O1 has been detected as a cause of cholera. Most of the non-O1 serotypes that persist in environmental water bodies are non-toxigenic and are considered normal inhabitants of environmental water.^5^ Consequently, detecting pandemic-like toxigenic *V. cholerae* O1 in environmental samples is confounded by the presence of related but non-epidemic strains.^5–7^

Standard methods for detecting *V. cholerae* in water samples generally begin with concentration of a relatively large volume of water, often by membrane filtration of approximately 500 ml through a Millipore filter.^8^ ^9^ This usually requires a pump or vacuum system. The filter is then incubated in alkaline peptone water (APW) for 6–18 hours to selectively enrich *V. cholerae*, followed by plating onto thiosulfate-citrate-bile salts-sucrose (TCBS) agar and overnight incubation. Presumptive yellow colonies are then subcultured onto nonselective media such as LB or nutrient agar for biochemical testing and confirmation by agglutination with O1 antiserum. Extensive supplies and filtering equipment, combined with at least seventy-two hours to obtain results, significantly constrain this gold standard procedure.

In this study, we developed a simpler, faster, and less expensive method for detecting *V. cholerae* O1 in environmental water samples. The approach uses gauze filtration instead of membrane filtration and detects *V. cholerae* directly from APW enrichment using an RDT. We aimed to create a method that is inexpensive, easy to deploy at multiple sites, and suitable for field-based surveillance. At the same time, the method needed to be compatible with culture and / or molecular confirmation, including PCR, to ensure performance comparable to established laboratory approaches. We therefore validated the assay using field samples naturally contaminated with *V. cholerae* in Lagos, Nigeria, a region where cholera is frequently reported.

We previously applied this method in Cameroon and Uganda during times when there were no cases occurring. In those studies, environmental samples were negative for *V. cholerae* O1, although many were positive for non-O1 *V. cholerae.*^10,11^ Testing samples from Nigeria provided an opportunity to further evaluate the method in a cholera-endemic setting and to examine environmental contamination in a country that experiences substantial cholera burden, with approximately 20,000 reported cases annually across many states.^12–14^ Lagos is particularly relevant because it experienced a major outbreak in 2024 that extended into 2025. The city lies on the Atlantic coast and is characterized by low-lying terrain, lagoons, canals, barrier islands, and peninsulas that are highly vulnerable to fecal contamination due to inadequate sewage infrastructure. Previous studies have detected a range of pathogens, including *V. cholerae*, in these waters.^15,16^

## Methods

During 2025, we conducted monthly surveillance for *V. cholerae* from surface water sites in Lagos, Nigeria. We collected one liter grab samples from each of the sites and filtered the water through a sterile gauze pad that was tightly fit into the spout of a funnel as described by Spira and Ahmed.^17^ We used clean plastic water bottles for collecting the water and used the same type of plastic water bottles as funnels after cutting away the base of the bottle. Once the water passed through the gauze pad, the pad which contains about 20 ml of water, was transferred into a 50 mL conical tube with 20 mL double strength APW. The APW was then incubated overnight at 37°C. The next morning, the APW was tested for *V. cholerae* O1 using an RDT (Crysal VC- O1) directly by placing four drops of the APW into the test tubes and inserting the test strip into the tube without using the buffer included with the kit. Two drops of the APW were also spotted onto each of two different spots on filter paper (Whatman 903 Protein Saver Cards) and the filter paper samples were placed in a protected area to dry at room temperature. When dry, the filter papers were placed in paper envelopes and then into a plastic bag and stored at room temperature until tested later by PCR. To isolate *V. cholerae* bacteria, samples of APW that were RDT positive were streaked onto TCBS agar and incubated overnight at 35° to 37° C. Typical yellow colonies were further identified as *V. cholerae* O1 using standard methods including agglutination with O1 antisera and serotype specific antisera. Isolates of *V. cholerae* from the TCBS were preserved on agar slants and frozen in 15% glycerol media. To preserve DNA from the isolates, the isolates were inoculated into APW and incubated for five hours, after which two drops were placed onto each of two spots on the filter paper and, as with the APW samples, were allowed to dry before placing in envelopes and baggies.

The filter paper samples from the overnight APW and the isolates were sent to Johns Hopkins University, where DNA was extracted from the filter paper via boiling extraction with Chelex^18^ and the DNA was tested for toxigenic V. cholerae O1 by conventional PCR targeting genes for ompW and ctxA. ^10,19^ ompW codes for an outer membrane protein that is found in all *V. cholerae* including non-O1 serotypes, while ctxA is specific for toxigenic *V. cholerae*. Samples that were positive for both ompW and ctxA were deemed to be toxigenic *V. cholerae* O1 while samples that were ompW positive, but ctxA negative were deemed to be non-toxigenic, non-O1 *V cholerae*.

In addition, to confirm the results of the conventional PCR and extend the findings to include additional genes, we tested the filter paper samples from the overnight APW using two different qPCR procedures. One assay is an in-house assay that targets ompW^20^, wbeO1^21^, and ctxA^22^ using the New England Biolabs Luna Universal Probe qPCR Master Mix (Cat# M3004X); the other is a commercial kit (Bioperfectus Reat Time PCR Kit (Jiangsu Bioperfectus Technologies Co., Ltd, Cat# YJB10401W) that targets ctxA, serotypes O1 and O139.

To confirm that this process did not alter the *V. cholerae* bacteria, we carried out whole genome sequencing of 20 RDT-positive samples (from APW) matched with the corresponding isolates stored on filter paper. We also compared the sequences to five recent anonymous clinical samples from Lagos (isolates from stool), to ensure the lineages found in the environmental samples adequately reflected clinical circulation. Sequencing libraries were generated from DNA extracts using the Illumina DNA Prep kit. Sequencing libraries from all samples were pooled equimolarly and sequenced on a single lane of a NovaSeq X 10B 2x150bp run. The samples received 29 million reads on average.

Sequencing data for each sample was analyzed using the TheiaProk Illumina paired-end sequencing workflow v4.2.0 on Terra.^23^ Briefly, the workflow performs de novo genome assembly, quality control, and characterization of bacterial sequencing data. The presence of antimicrobial resistance genes was determined with AMRFinderPlus v4.2.7 using database version 2026-03-24.1.^24^ We used Vibecheck (https://github.com/CholGen/Vibecheck) to classify samples as canonical transmission event lineages (i.e., AFR1-AFR17), using raw reads as input. These methods are illustrated in Figure 1.

**Figure 1.**
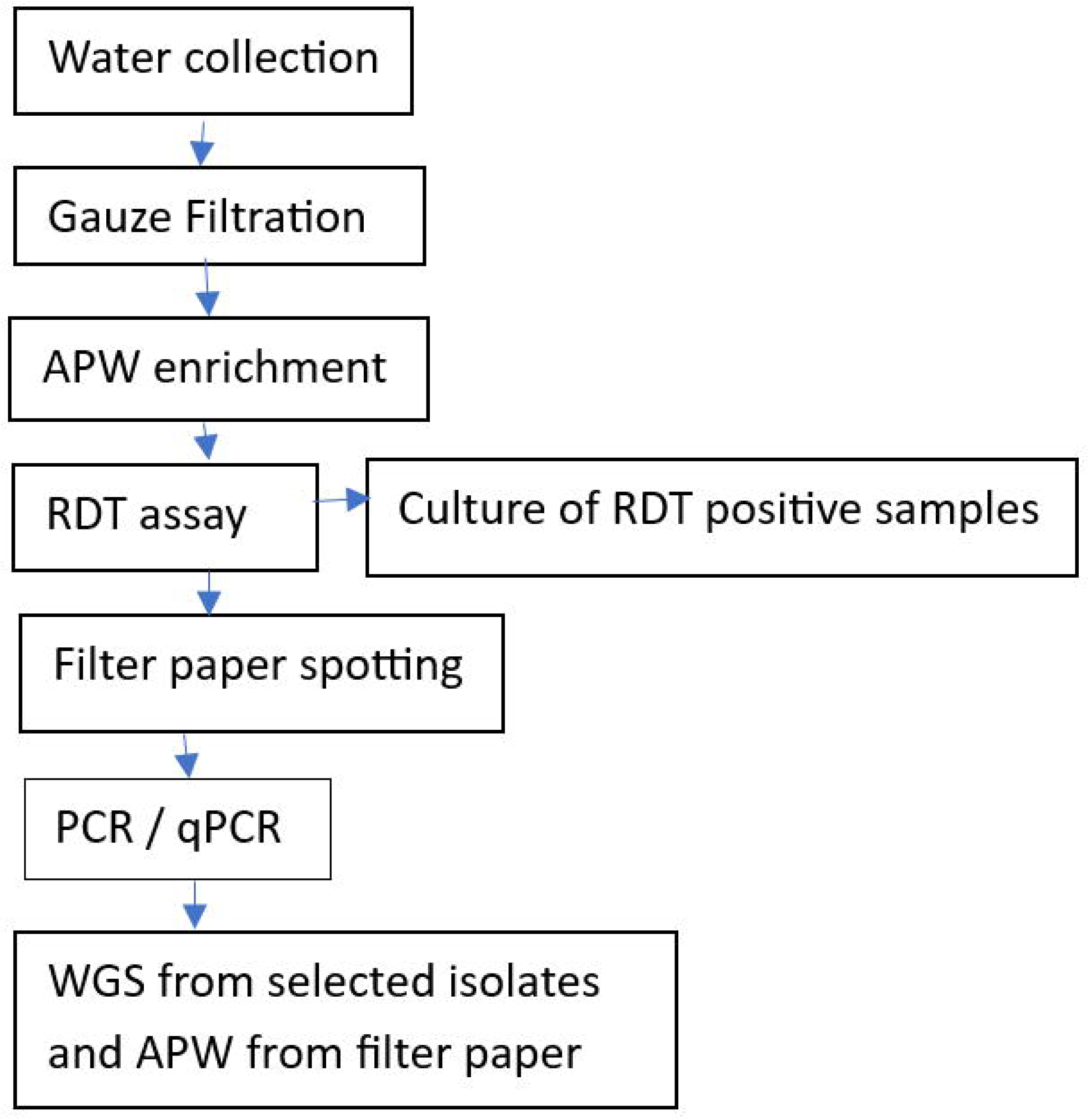
Flow chart of the procedures to detect and validate *V cholerae* O1 from environmental water samples in Lagos Nigeria.

## Results

Between 12 March and 21 October 2025, we collected and tested ninety-five water samples from seventeen sites in Lagos, Nigeria using this simplified method. The overall RDT results and their agreement with the conventional and qPCR assays are shown on Table 1.

**Table 1.** Results of environmental water samples that were tested with rapid diagnostic tests and PCR assays following incubation in APW in Lagos Nigeria.

|  | PCR |  |  | qPCR (both assays) |  |  | Bioperfectus qPCR |
| --- | --- | --- | --- | --- | --- | --- | --- |
|  | *ompW*<br>pos | *ctxA*<br>pos | Neg for<br>both | *ompW*<br>pos | *ctxA*<br>pos | O1 pos | O139 pos |
| RDT pos | 54 | 54 | 0 | 54 | 54 | 54 | 0 |
| RDT neg | 24 | 0 | 17 | 24 | 0 | 0 | 11 |

Fifty-four samples were RDT positive and these were also culture positive. They were also confirmed to be positive by PCR for *omp*W and *ctx*A by both conventional and qPCR. As a result, all were deemed to be toxigenic *V. cholerae* O1. Twenty-four samples were RDT negative but were positive for *ompW* but negative for *ctxA* by both conventional and qPCR. These were deemed to contain non-toxigenic, non-O1 *V. cholerae.* Among the sites with non-O1 *V. cholerae*, eleven were positive for O139 serotype by qPCR using the Bioperfectus kit.

The locations of the sites and the types of *V. cholerae* identified are shown in Figure 2.

**Figure 2.**
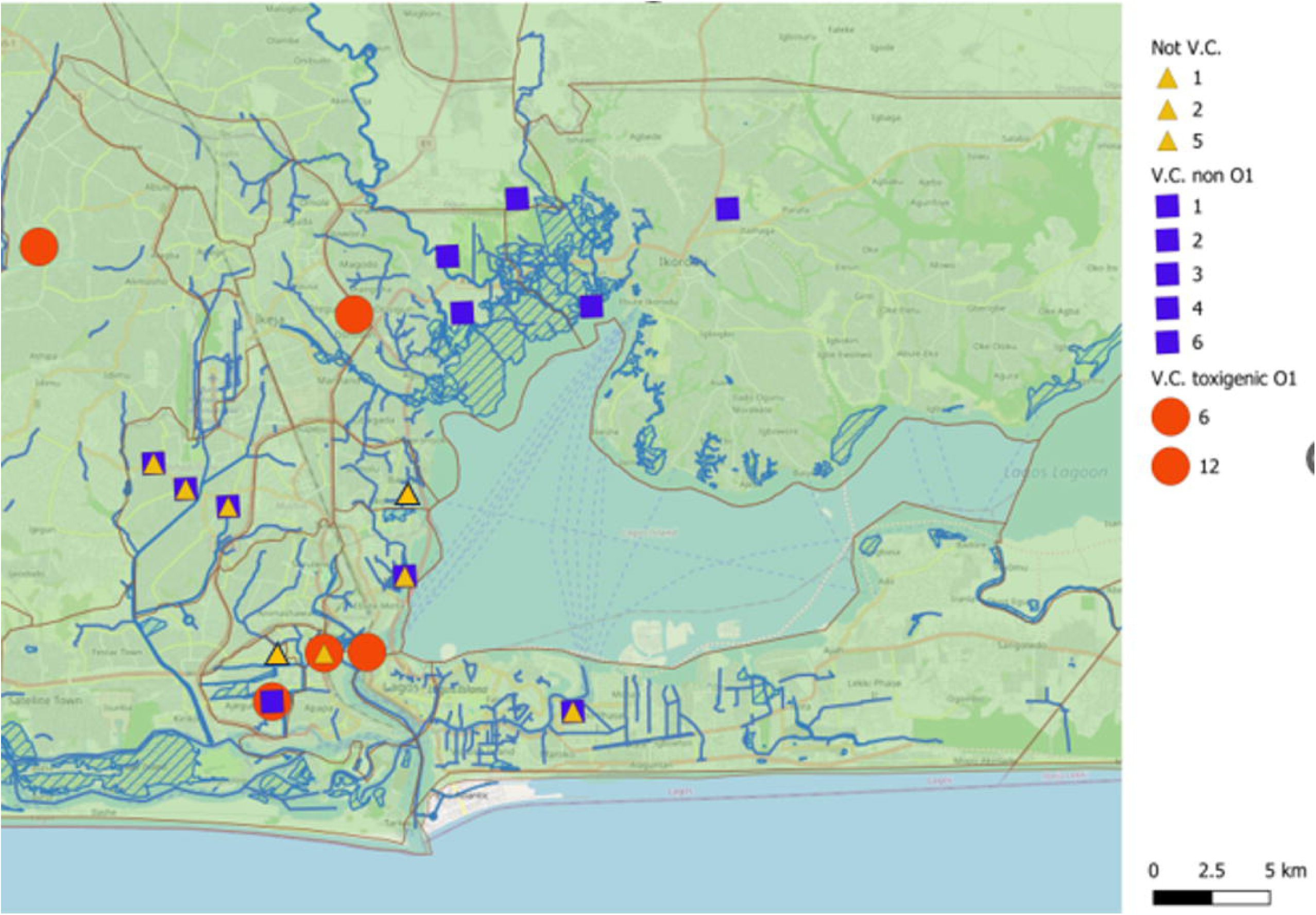
Map of Lagos, Nigeria showing sampling sites where *V. cholerae* was detected.

Of the seventeen sites, all the ctxA positive samples came from one of five of the sites (Iddo, Ijora, Ajegunle, Toll Gate and Aboru). Of these sites, four had twelve positive samples, and one (Ajegunle) had six samples that were positive for O1 *V. cholerae* and six that were positive for O139 *V. cholerae*. At this site, the O1 and O139 samples were collected at different time points, not concurrently. The other sites were negative by RDT. Figure 3 shows the times during the year when these different strains of *V. cholerae* were detected.

**Figure 3.**
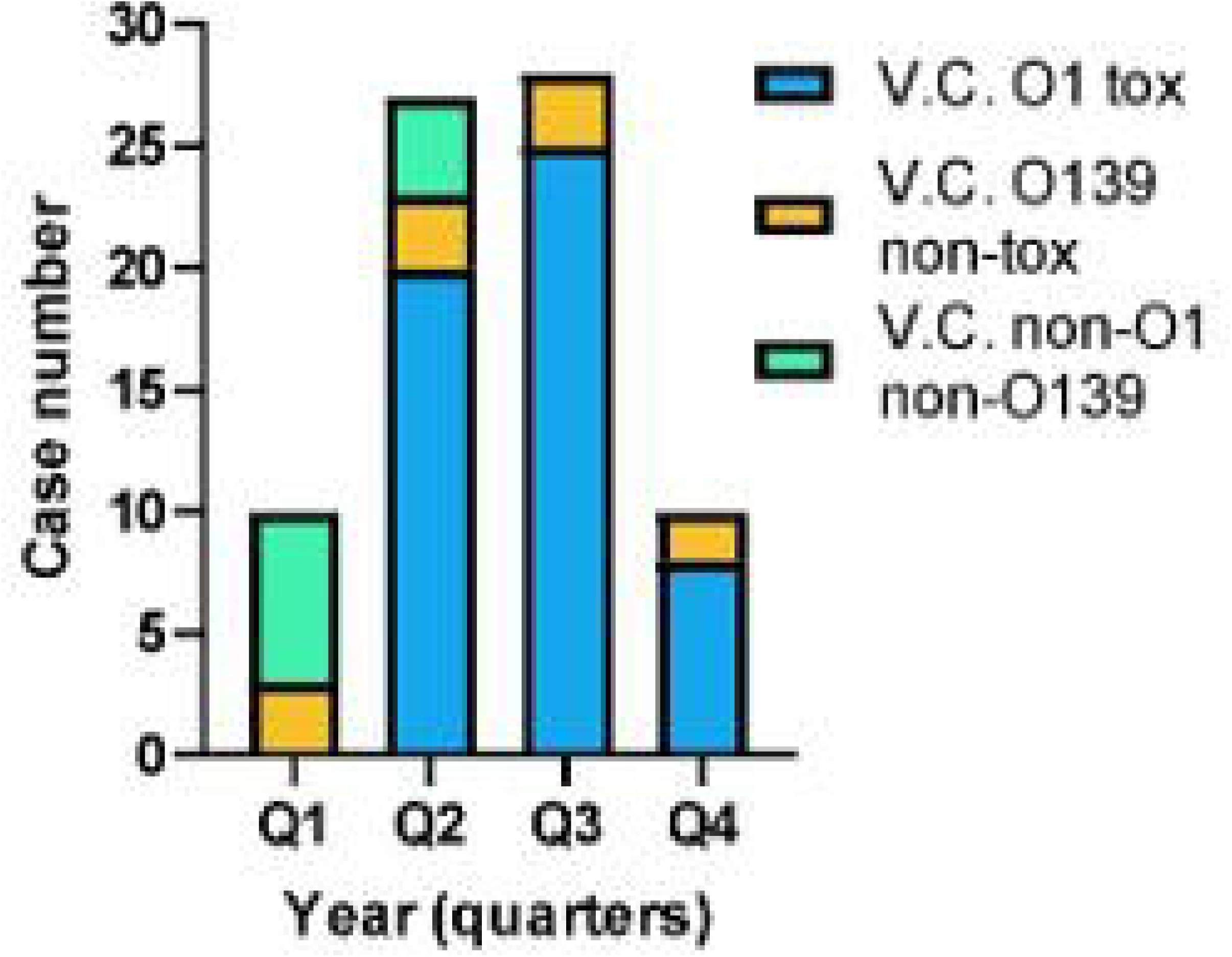
The graph shows the number of samples with toxigenic *V. cholerae* O1, non-toxigenic O139 *V. cholerae* and samples with non-toxigenic nonO1-non O139 *V. cholerae*. during the four quarters of 2025 in Lagos Nigeria.

Since the results from the initial RDT perfectly aligned with the PCR results, we determined that the RDT result following APW enrichment was sufficient to detect toxigenic *V. cholerae* O1 and estimated the costs for cholera surveillance using only the RDT following APW enrichment. The materials needed included plastic water bottles, gauze pads, 50 ml conical tubes, 20 ml of double strength APW and the RDT kits. The cost for these totaled less than $2.50 per sample, with the major costs being the conical tubes and the RDTs. Since the RDTs are now being provided through GAVI, the cost to the country will be lower.

We did WGS from twenty matched APW and environmental sample isolates from these water bodies as well as five of the clinical isolates to confirm that our method preserved DNA sufficiently for WGS. We found that we were able to generate high-quality sequencing results from most of the samples. For 15 matched pairs, WGS were available from both the APW sample and the isolate sample from the same water sample and the sequencing results were concordant with our expectations; specifically, all samples were AFR12, which is the primary lineage circulating in Nigeria since at least 2009.^22^ When comparing the WGS of the matched environmental APW and isolate samples from the same water sample, more antibiotic resistance genes were detected in the APW samples than the matching isolate samples from the same water body.

Commonly detected resistance genes across the samples include almE, almF, almG, varG, floR, gyrA, sul1, sul2, tet(A), tet(B), catB9, drfA1, drfA15, APH(3”)-lb, and APH(6)-ld.

## Discussion

This efficient, low-cost method of detecting *V. cholerae* O1 from contaminated environmental water provided reliable results within one day of sample collection. All of the RDT positive samples yielded *V. cholerae* O1 isolates when cultured on TCBS, and the RDTs results matched the results from conventional PCR and qPCR. By comparison, standard methods which involve Millipore filtration that relies on identifying *V. cholerae* by culture require at least three days and also require more technical expertise. We have used RDTs following enrichment in APW previously^10,25,26^; however, this is the first use of this method to combine gauze filtration with RDT detection and confirmation with PCR and culture from field samples from an endemic area.

There are several advantages to this method. By quickly identifying the contaminated water source, interventions could theoretically be quickly implemented to prevent people from using this water source without treating or boiling. Secondly, because of the simplicity of the procedure, samples can be collected from multiple sites, allowing for screening to take place over multiple sites and times. This facilitates identification of contaminated water bodies and enables epidemiological assessments of risks. Thirdly, detection of *V. cholerae* O1 from water may detect human infections that are not apparent from clinical surveillance alone. Finally, when one wishes to isolate *V. cholerae* from environmental water for further study, the RDT results can screen samples to identify the samples most likely to contain *V, cholerae;* thus, avoiding the need to culture so many samples that are likely to be negative.

The map showing the locations of the water bodies sampled suggest a clustering with the same site being positive for toxigenic *V. cholerae* multiple times. We did not explore possible reasons for this clustering, but this experience does suggest that further epidemiologic studies can be undertaken to understand the source of the contamination and the factors leading to its persistence. With results available so quickly, one could potentially sample surrounding areas or upstream to identify the true source of the contamination.

We found that the filter paper samples from APW enrichment and isolates were suitable for WGS. Other studies have shown the stability of nucleic acid on filter paper^27^. The availability of these alternative samples for WGS makes possible the studies of cholera from remote areas where culture is not feasible. However, the APW samples contain nucleic acid from other microorganisms and detection of AMR genes from APW samples will be confounded by the presence of these other species.

Interestingly, most of the samples positive for toxigenic *V. cholerae* were identified during the rainy season (April through September); however, the samples positive for non-O1 were seen throughout the year. Also of note, some of the sites found positive for toxigenic *V. cholerae* also yielded samples positive for non-O1 *V. cholerae*. Our methods, however, do not allow us to determine if the two might be occurring simultaneously. When RDT positive samples were cultured, the isolates identified on TCBS media were found to be toxigenic *V. cholerae* O1. This suggests that the toxigenic strains were predominant; if non-O1 strains were present, they were likely in lower concentrations.

Some limitations from our study should be mentioned. This study used Crystal VC O1 which is an RDT designed to detect only serotype O1. This could be considered an advantage since serotype O1 is the serotype of importance and interest, but it should be noted that the more commonly used RDTs include lines for both serotypes O1 and O139. Since the Bioperfectus qPCR results suggest the presence of non- toxigenic *V. cholerae* O139 at some of these water sites, it is possible that the RDTs with lines for both serotypes might detect *V. cholerae* O139. However, a positive line for O139 may also be confusing when one intends to detect only serotype O1.

A second limitation relates to the possibility that the test might not detect very low concentrations of *V. cholerae* in environmental water samples. The use of gauze filtration rather than Millipore filtration is based on the association of the bacteria adhering to larger particles such as zooplankton and phytoplankton that would be retained in the gauze and be amplified in the APW.^28^ *V. cholerae* bacteria that are not attached to these larger particles might flow through the gauze filter and may not be captured. Further studies may need to be carried out comparing the standard methods using a Millipore filter with this gauze filtration method.

Clearly, this is a qualitative and not a quantitative assay. Others have used a quantitative assay to determine the degree of risk from use of environmental water from directly drinking the water.^29^ We did not attempt to determine this risk since any water containing *V. cholerae* O1 should always be considered risky. Even low numbers of bacteria can multiply when introduced into food. ^30^

## Conclusion

The use of RDTs after APW enrichment using gauze filtration was found to be a reliable method to detect *V. cholerae* O1 from environmental water. This simplified method was found to be convenient, inexpensive, and provided results within a day. It should serve as a practical tool to monitor cholera transmission and persistence in environmental water samples. Several applications for this procedure include epidemiological surveillance, outbreak investigation, and municipal water supply monitoring. Future studies of environmental water may also choose to include detection of vibriophage as an additional maker of *V cholerae* contamination. Recently we published a simple and convenient method for detecting these phages, and a procedure for detecting both the bacteria and the phage could be developed.^31^

## Supporting Information

## Acknowledgments

We thank the field workers in Lagos who collected the samples.

## Address for Correspondence

David A Sack, M.D. Professor, Department of International Health, Johns Hopkins Bloomberg School of Public Health, 615 North Wolfe Street, E5537, Baltimore, Maryland 21205, USA., +1 443 287 8795

## Data Availability

Data can be made available upon request.

